# Measuring and Reducing Racial Bias in a Pediatric Urinary Tract Infection Model

**DOI:** 10.1101/2023.09.18.23295660

**Authors:** Joshua W. Anderson, Nader Shaikh, Shyam Visweswaran

## Abstract

Clinical predictive models that include race as a predictor have the potential to exacerbate disparities in healthcare. Such models can be respecified to exclude race or optimized to reduce racial bias. We investigated the impact of such respecifications in a predictive model – UTICalc – which was designed to reduce catheterizations in young children with suspected urinary tract infections. To reduce racial bias, race was removed from the UTICalc logistic regression model and replaced with two new features. We compared the two versions of UTICalc using fairness and predictive performance metrics to understand the effects on racial bias. In addition, we derived three new models for UTICalc to specifically improve racial fairness. Our results show that, as predicted by previously described impossibility results, fairness cannot be simultaneously improved on all fairness metrics, and model respecification may improve racial fairness but decrease overall predictive performance.

## Introduction

The healthcare community is gaining interest in the investigation of bias in predictive models that are derived using statistical and machine learning (ML) methods. Model biases can lead to decisions that cause unfairness or discrimination against specific groups of people. As is common in the fairness literature, we will use the terms bias, unfairness, and discrimination, interchangeably [1]. The use of predictive models to aid in clinical decision-making is increasing, and some models include race as a predictor because it may be associated with the outcome being predicted. However, it is becoming increasingly clear that the physical trait of skin color, which is used to define race, is a poor proxy for biological or genetic variation among individuals. Despite the lack of a meaningful correspondence between race and biology, race is frequently used in medical research to explain disease prevalence and outcomes, as well as in clinical medicine for decision making [2]. Clinical predictive models that use race as a predictor may guide clinical decisions in ways that favor individuals of racial majorities over members of ethnic minorities. In such models, it is critical to investigate whether and how race is an important predictor of the outcome and the implications of including or excluding it. Despite increased awareness of model bias, research in bias of clinical predictive models is still in its infancy [3]. For instance, the well-established link between urinary tract infection (UTI) and race prompted the development of a logistic regression model, UTICalc, for predicting pediatric UTI that included race as a predictor [4]. The inclusion of race in the model was subsequently reconsidered and it was replaced with alternative predictors [5].

An unfair clinical decision due to the guidance of a statistical or ML model results in unexpected favoritism or prejudice towards a specific group or groups [6]. In this study, we assess the fairness of a clinical predictive model, UTICalc, which is designed to assist clinicians in identifying children likely to need collection of urine by urethral catheterization to definitively establish UTI. We examine whether the revised model reduced racial discrimination relative to the original model, and whether ML methods can be applied to such existing clinical models to increase fairness. Using a variety of fairness metrics, we quantify the fairness of UTICalc across black and nonblack children. In addition, we investigate if racial fairness of UTICalc could be improved by optimizing the logistic regression coefficients, incorporating additional predictors in the model, or both.

## Background

### Clinical models and race

Models that predict clinical outcomes, also known as clinical algorithms, are increasingly integrated into electronic health records, guidelines, and decision support tools, and there is concern that using models that include race as a predictor may result in disparities in health care, particularly among racial minority populations. Using race as a proxy for genetic or broader biological differences can oversimplify complex health issues and contribute to disparities. It is now widely acknowledged that race is a historical and social construct, not a biological one. The use of race in clinical decision-making has come under increasing scrutiny as it has become more apparent that race-based diagnosis and treatment reflect flawed biological and genetic assumptions. Many models that include race as a predictor are in clinical use. A recent article identified 39 risk calculators that use race as a predictor, six laboratory test results with different reference ranges recommended for different races, one therapy recommendation based on race, and fifteen medications with initiation and monitoring guidelines based on race [7]. However, it is unclear whether the inclusion of race as a predictor to inform clinical decision-making will always perpetuate long-standing disparities in health and healthcare [8]. Therefore, eliminating race is a more nuanced process than simply removing it from a clinical model, and the implications of removing race from a model should be quantified.

### Model fairness

Model fairness, more commonly known as algorithmic fairness, refers to the notion that a statistical or ML model should predict outcomes such that they lead to fair and equitable decisions [2]. This is becoming increasingly important as models are used in a wide range of decision-making, such as in job recruitment, criminal sentencing, and medicine. In the context of race, the goal of model fairness is to ensure that outcome predictions are not racially biased or discriminatory. Model fairness is not driven by model performance [9]. Models that have strong overall performance can still have large disparities in performance across racial groups [2].

Research on bias, unfairness, and discrimination has a long history in several domains [10], which predates research on fairness in clinical models. Researchers have proposed several formal definitions of model fairness for assessing bias. There are two broad categories of model fairness definitions: group fairness and individual fairness. The concept of group fairness states that a model’s predictions should not result in discrimination based on membership in specific groups of the population. Such socially salient groups include those who have historically faced systematic discrimination; for example, federal laws in the United States recognize protected groups based on race, color, sex, age, disability status, and national origin. In a model, a feature that encodes explicitly or implicitly an individual’s status in a protected group is called a sensitive attribute. The objective of group fairness is to ensure that the rate of a particular outcome (e.g., being hired, being sentenced to prison, being catheterized for obtaining a urine sample) is roughly the same across groups defined by a sensitive attribute. Group fairness partitions the population on a sensitive attribute (e.g., race) and imposes outcome invariance on the groups (e.g., black and nonblack groups). In this study, we focus on group fairness because it lends itself to statistical analysis and is the subject of most prior research.

Individual fairness focuses on the idea that a model should generate similar predictions for individuals who are similar in ways that are relevant to the decision at hand. The similar treatment principle, which states that similar people should be treated similarly, embodies this idea of fairness [11]. Individual fairness frequently employs two distance metrics: the first measures how similar any two individuals are, and the second measures how similarly those individuals are treated. Individual fairness requires that if two individuals are close on the similarity metric, they should also be close on the treatment metric. In the literature, comparatively fewer studies have investigated individual fairness than group fairness.

## Sources of bias

In the literature, numerous causes of unfairness have been identified [12] including biases in problem specification, data, modeling, and deployment [13].

### Biases in problem specification

The development of a predictive model necessitates specifying the overall goals, actions available for decision making, and, in the case of predicting an outcome of interest, defining well an outcome that is likely to be complex and ambiguous. For example, Obermeyers et al. demonstrated that a healthcare model used to identify patients with complex health needs was racially biased because the outcome was specified using healthcare costs as a proxy for illness burden. Because of unequal access to healthcare, the model unfairly predicted that members of groups with less access were healthier than they were [14].

### Biases in data

Following problem specification, model derivation typically entails applying a statistical or ML method to a dataset of historical cases in order to discover useful patterns. Biases in the data will be captured in the model because model derivation methods aim to capture statistical features of the input dataset. For example, if there is systemic racism in healthcare access and delivery, historical EHR data will encode these biases and lead to discriminative predictive models [15].

### Biases in modeling

Given a dataset, derivation and validation of a model is done to optimize model performance relative to some criteria of success which is technically encoded as an objective function. Typically, the objective function focuses on pure overall predictive performance by maximizing accuracy. Since fairness is not explicitly accounted for in the objective function, such optimization may lead to models that have lower accuracy in a specific group or groups.

### Biases in deployment

Finally, biases can emerge during the deployment of a model. This can occur, for example, when the deployment context differs significantly from the historical context of the training data, resulting in poor performance in groups that were underrepresented in the training data. Furthermore, biases can arise from a failure to recognize the limitations of a purely predictive model, whose predictions may be insufficient for interventions that require causal information.

## Fairness metrics

Many of the metrics that are used to evaluate the performance of statistical and ML models have been adapted for measuring group fairness. Broadly speaking, group fairness metrics can be categorized into metrics of discrimination, calibration, and proper scoring rules. Discrimination metrics evaluate how well a model differentiates among the various classes or values of the target variable. Calibration is the concordance of predicted probabilities with the occurrence of positive instances. Proper scoring rules assess the quality of probability predictions and evaluate both discrimination and calibration of a model. Table 1 lists the fairness metrics we used in our experiments and includes 11 discrimination metrics, one calibration metric (expected calibration error) and one proper scoring rule (Brier score).

**Table 1.** Definitions of fairness metrics.

| Fairness metric | Definition | Description in our study |
| --- | --- | --- |
| Demographic parity (DP) | The outcomes are equal:<br>$P(\hat{Y} = 1 \vert R = b) =$<br>$P(\hat{Y} = 1 \vert R = nb)$ | The rates of predicted UTI are equal in both racial groups. |
| Equal opportunity (EOP) | The true positive rates (TPRs) are equal:<br>$P(\hat{Y} = 1 \vert R = b, Y = 1) =$<br>$P(\hat{Y} = 1 \vert R = nb, Y = 1)$ | The TPRs are equal in both racial groups. TPR is the proportion of those who have UTI (true positives + false negatives) and are correctly predicted to have UTI (true positives). |
| Specificity (Spec) parity | The true negative rates (TNRs) are equal:<br>$P(\hat{Y} = 0 \vert R = b, Y = 0) =$<br>$P(\hat{Y} = 0 \vert R = nb, Y = 0)$ | The TNRs are equal in both racial groups. TNR is the proportion of those not have UTI (true negatives + false positives) and are correctly predicted not to have UTI (true negatives). |
| False positive rate (FPR) parity | The FPRs are equal:<br>$P(\hat{Y} = 1 \vert R = b, Y = 0) =$<br>$P(\hat{Y} = 1 \vert R = nb, Y = 0)$ | The FPRs are equal in both racial groups. FPR is the proportion of those who do not have UTI (true negatives + false positives) and are wrongly predicted to have UTI (false positives). |
| False negative rate (FNR) parity | The FNRs are equal:<br>$P(\hat{Y} = 0 \vert R = b, Y = 1) =$<br>$P(\hat{Y} = 0 \vert R = nb, Y = 1)$ | The FNRs are equal in both racial groups. FNR is the proportion of those who have UTI (true positives + false negatives) and are wrongly predicted to not have UTI (false negatives). |
| Equalized odds (EOD) | The odds are equal if $\hat{Y} \perp\!\!\!\perp R \vert Y$:<br>$P(\hat{Y} = 1 \vert R = b, Y = y) =$<br>$P(\hat{Y} = 1 \vert R = nb, Y = y)$<br>where $y \in \{0, 1\}$ | The TPRs and FPRs are equal in both racial groups. |
| Positive predictive value (PPV) parity | The PPVs are equal:<br>$P(Y = 1 \vert R = b, \hat{Y} = 1) =$<br>$P(Y = 1 \vert R = nb, \hat{Y} = 1)$ | The PPVs are equal in both racial groups. PPV is the proportion of those predicted to have UTI (true positives + false positives) and do have UTI (true positives). |
| Negative predictive value (NPV) parity | The NPVs are equal:<br>$P(Y = 0 \vert R = b, \hat{Y} = 0) =$<br>$P(Y = 0 \vert R = nb, \hat{Y} = 0)$ | The NPVs are equal in both racial groups. NPV is the proportion of those predicted to not have UTI (true negatives + false negatives) and do not have UTI (true negatives). |
| Accuracy (Acc) parity | The accuracies are equal:<br>$P(\hat{Y} = y \vert R = b, Y = y) =$<br>$P(\hat{Y} = y \vert R = nb, Y = y)$<br>where $y \in \{0, 1\}$ | The accuracies are equal in both racial groups. |
| Matthew's correlation coefficient (MCC) parity | The MCCs are equal:<br>$P(MCC(Y, \hat{Y}) \vert R = b) =$<br>$P(MCC(Y, \hat{Y}) \vert R = nb)$ | The MCCs are equal in both racial groups. MCC measures the differences between actual and predicted values and is equivalent to the chi-square statistic for a two-by-two contingency table. |
| Area Under the ROC curve (AUROC) parity | The AUROCs are equal:<br>$P(AUROC(Y, \hat{Y}) \vert R = b) =$<br>$P(AUROC(Y, \hat{Y}) \vert R = nb)$ | The AUROCs are equal in both racial groups. AUROC is commonly used to measure how well a model discriminates between two classes. |
| Expected calibration error (ECE) parity | The ECEs are equal:<br>$P(\hat{Y} \vert R = b) = P(Y \vert R = b),$<br>$P(\hat{Y} \vert R = nb) = P(Y \vert R = nb)$ | ECE is equal in both racial groups. ECE measures how closely the estimated probabilities of a model correspond to the actual probabilities. |
| Brier score (BS) parity | BSs are equal:<br>$\frac{1}{N} \sum_{i=1}^N (\hat{Y}_i \vert R = b - Y_i \vert R = b)^2 = \frac{1}{N} \sum_{i=1}^N (\hat{Y}_i \vert R = nb - Y_i \vert R = nb)^2$ | BSs are equal in both racial groups. BS is a proper scoring method measuring accuracy of predictions. |

$Y$ represents the outcome (UTI), $\hat{Y}$ represents the prediction (prediction of UTI), and race is represented as $R$ with values $b$ for "black" and $nb$ for "nonblack."

Demographic parity (DP), also known as statistical parity, requires that the proportion of positive predictions is the same across all groups defined by a sensitive attribute [2]. Since DP does not assess the quality of the predictions, imposing DP may conceal harmful impacts. Accuracy (Acc) parity requires equal accuracy across groups. While Acc parity addresses some of the shortcomings of DP, it does not differentiate between error types. Acc parity is appropriate when the effects of false positives and false negatives are comparable.

Equal opportunity (EOP) requires that the sensitivity or true positive rate (TPR) be the same for all groups and specificity (Spec) parity requires that the specificity or true negative rate (TNR) be the same for all groups. When both EOP and Spec parity are satisfied, equalized odds (EOD) is satisfied. False positive rate (FPR) and false negative rate (FNR) parity require that FPR and FNR be the same for all groups.

Equalized odds (EOD) is satisfied when both EOP and Spec parity are satisfied. False positive rate (FPR) parity and false negative rate (FNR) parity require FPR and FNR to be the same for all groups, respectively. Positive predictive value (PPV) parity requires that the PPV be the same for all groups and negative predictive value (NPV) parity requires that the NPV be the same for all groups. Predictive value parity is satisfied when both PPV parity and NPV parity are satisfied. Matthew’s correlation coefficient (MCC) measures the differences between actual and predicted values and is equivalent to the chi-square statistic for a two-by-two contingency table. MCC is a more constraining metric than Acc, as it requires a high TPR and FPR to obtain a high MCC [16]. MCC parity is satisfied when MCC is the same for all groups. Area under the receiver operating characteristic curve (AUROC) is commonly used to measure how well a model discriminates between two classes. AUROC parity requires that the AUROC be the same for all groups.

Expected calibration error (ECE) is a weighted average of the accuracy versus prediction errors across bins where the bins are created by dividing the probability interval into multiple bins. The ECE measures how closely the estimated probabilities of a model correspond to the actual probabilities. ECE parity requires that the model be calibrated equally across all groups. Brier score (BS) is a proper scoring method measuring accuracy of predictions. For predicted probabilities, BS is equivalent of mean squared error (MSE). BS parity requires the BS is the same for all groups.

### Independence, separation, and sufficiency

The large array of fairness metrics described above relate to the three main broad notions of independence, separation, and sufficiency [17]. While the independence criterion relies only on the sensitive variable *R* and the prediction *Ŷ*, separation and sufficiency criteria also assess on error rates, thus relying on the outcome variable *Y* as well. Independence asserts that the prediction is independent of the sensitive feature which in our case is race, i.e., (*Ŷ* ╨ *R*). In general, we expect some correlation between race and the other predictors [18]. As a result, we cannot guarantee fairness based on independence by simply removing race as a predictor; rather, the model must treat the groups differently. Separation asserts that the prediction is independent of race conditioned on the actual outcome, i.e., (*Ŷ* ╨ *R* | *Y*) [19]. For more details, see the definition of equalized odds in Table 1. This implies that if the outcome is correlated with race, there is a possibility of bias even with a difference of 0 [17]. Sufficiency asserts that the actual outcome is independent of race conditioned on the prediction, i.e., (*Y* ╨ *R*|*Ŷ*) [20]. For more details, see the definition of positive predictive value parity in Table 1. This specifically focuses on parity between groups with the same outcomes. Thus, the metrics we included capture these different notions of fairness.

### Impossibility results

Given the variety of fairness measures and parity conditions, one question is whether all of these requirements can be met concurrently. It turns out that due to the conflicting notions of fairness embodied in the metrics, it is impossible to optimize a model on all metrics. Consider a predictive model *M* (*X, R*) that predicts a binary outcome *Y* from predictors *X* and a sensitive variable *R* with an unequal prevalence of *Y* across *R*. Then some impossibility results include:

- If *R* and *Y* are not independent, then DP and predictive value parity cannot simultaneously hold.
- If *R* and M are not independent of *Y*, then DP and EOD cannot simultaneously hold.
- If *R* and *Y* are not independent, then EOD and predictive value parity cannot simultaneously hold.

These impossibility results hold in the real world because predictive models are imperfect, the prevalence of outcomes is rarely equal across groups, *R* and *Y* are frequently associated, and *M* and *Y* are typically associated when the model performs well. Additional impossibility results have been described. For example, ECE parity and EOD cannot simultaneously be satisfied unless the model is perfect and is always accurate [21].

### UTICalc model

In children younger than 2 years, for which the UTICalc model was developed, obtaining a urine sample is challenging, and a clinical tool that predicts the probability of UTI from clinical features that are readily obtained in the emergency department is useful in identifying children who may benefit from urine sampling testing. UTICalc Version 1 (UTICalc v1), developed in 2018, is a logistic regression (LR) model that predicts the risk of UTI in febrile children 2 years of age or younger from five binary demographic and clinical predictors including age (*<*12 months vs ≥2 months), sex (female or uncircumcised male vs circumcised male), race (black vs nonblack), fever (*<*39°C vs ≥39°C), and alternate fever source (no other source of fever vs other source of fever) [5]. The authors of UTICalc v1 reasoned that as a screening tool the the model should achieve a minimum of 95% sensitivity for clinical use [4]. With a 2% cutoff, the model achieved a 95% sensitivity with the training data [5]. To mitigate the potential bias arising from including race as a predictor, in 2022, UTICalc Version 3 (UTICalc v3) was developed. UTICalc v3 is an updated LR model which was developed by replacing race in UTICalc v1 with two new predictors including history of UTI (yes vs no) and duration of fever (*<*48 hours vs ≥48 hours). The overall accuracy of the new model was equivalent to that of the initial model [22].

## Materials and Methods

### Dataset

The UTICalc dataset consists of data on 1,686 children and includes 11 features and one outcome variable that were extracted from EHR data at a single institution. In addition to the original model features, if urine analysis was available, additional features included nitrate was positive (yes vs no), bacteria was present on gram stain (yes vs no), white blood cell count (real), and leukocyte esterase (none, trace, 1-2, 2-3, or 3+) [4]. The outcome variable is the presence of UTI (yes vs no). Due to missing values, the sample sizes used to derive UTICalc v1 and v3 are different. UTICalc v1 used data from 1,593 children (407 blacks and 1,186 nonblacks) and UTICalc v3 used data from 1,552 children (398 blacks and 1,154 nonblacks).

### Deriving models to optimize fairness

We derived three new LR models to improve fairness across the black and nonblack groups. Two of them were derived using the exponentiated gradient reduction method that uses costs associated with predicting UTI for each patient [23]. Costs are derived for each patient using known methods [24]. Using normalized Lagrangian multipliers, this method solves a constrained optimization problem [23]. In simple terms, features of the model are given weights rather than assuming uniform importance.

In our first new model (UTICalc v1 Reweighted), we used the predictors from UTICalc v1 and reweighted the feature coefficients using exponentiated gradient reduction. Because UTICalc was designed to be a screening tool, the emphasis is on high sensitivity [5], and we focused our efforts on improving fairness of EOP that equalizes sensitivity values in the two racial groups. In the second new model (UTICalc All Features), all eleven predictors available in the dataset, including race, were used to derive a LR model. Because UTICalc v1 and UTICalc v3 have different specifications and, as a result, contain missing data, we used data from 1,171 children (298 blacks and 873 nonblacks). For the third new model (UTICalc All Features Reweighted), we used the predictors from UTICalc All Features and reweighted the feature coefficients using exponentiated gradient reduction. We used Microsoft’s Fairlearn (v0.8.0) package to perform exponentiated gradient reduction and the scikit-learn’s (v1.2.2) LogisticRegression method to derive the UTICalc All Features model.

### Statistical testing of fairness metrics

We measured fairness across racial groups with 13 fairness metrics. Each metric compares the values of a model performance metric between the black and nonblack groups (see Table 1). For comparisons, differences and ratios can be computed using the following formulae:

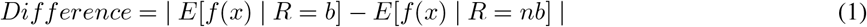

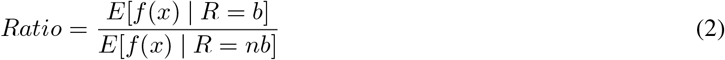

where *f* (*x*) is a fairness metric, *R* is the race predictor that takes values *b* (blacks) and *nb* (nonblacks). In our experiments, we used the difference, and assessed statistical significance of the difference between two models using the Wilcoxon ranked-sum test in bootstrapped distributions. Statistical significance was evaluated only between UTICalc v1 and UTICalc v3 models. A perfectly fair model will have exactly equal values for a particular fairness metric within each racial group, and hence the difference will equal zero.

### Model performance

In addition to the fairness metrics, we also report the predictive performance of the models for the black and nonblack groups. The performance metrics included AUROC, accuracy, sensitivity, and specificity. We thresholded the probability predictions at 0.5 to compute accuracy, sensitivity, and specificity.

## Results

Figure 1 shows the differences between 13 fairness metrics of the black and nonblack groups for five models and Table 2 provides the predictive performance of the five models on the same racial groups. UTICalc v1 had a large disparity in sensitivity between the racial groups as shown in Table 2. Mitigation methods varied widely in the predictive performance metrics. The exponentiated gradient method significantly worsened the UTICalc v1 Reweighted model’s sensitivity in both racial groups while improving accuracy and specificity. The UTICalc All Features model improved on all predictive performance metrics in the nonblack group but had mixed results in the black group, resulting in increased disparities compared to UTICalc v1 with the exception of sensitivity. The UTICalc All Features Reweighted UTICalc Reweighted had sensitivities above 90% and improved on other predictive performance metrics compared to UTICalc v1. UTICalc v3 significantly enhanced sensitivity in the black group while maintaining a high level of sensitivity in the nonblack group. It decreased specificity within the black group while increasing it within the non-black group.

**Table 2.** Predictive performance of the five models in the two the racial groups.

|  | **AUROC** | **Accuracy** | **Sensitivity** | **Specificity** |
| --- | --- | --- | --- | --- |
| **UTICalc v1** |  |  |  |  |
| Black (N=407) | 0.679 | 0.592 | 0.818 | 0.539 |
| Nonblack (N=1186) | 0.620 | 0.536 | 0.975 | 0.264 |
| **UTICalc v1 Reweighted** |  |  |  |  |
| Black (N=407) | 0.599 | 0.762 | 0.338 | 0.861 |
| Nonblack (N=1186) | 0.676 | 0.732 | 0.438 | 0.914 |
| **UTICalc All Features** |  |  |  |  |
| Black (N=298) | 0.867 | 0.809 | 0.969 | 0.969 |
| Nonblack (N=873) | 0.692 | 0.632 | 1.000 | 0.398 |
| **UTICalc All Features Reweighted** |  |  |  |  |
| Black (N=298) | 0.940 | 0.960 | 0.906 | 0.974 |
| Nonblack (N=873) | 0.960 | 0.964 | 0.938 | 0.981 |
| **UTICalc v3** |  |  |  |  |
| Black (N=398) | 0.729 | 0.455 | 0.935 | 0.340 |
| Nonblack (N=1154) | 0.810 | 0.590 | 0.962 | 0.358 |

**Figure 1.**
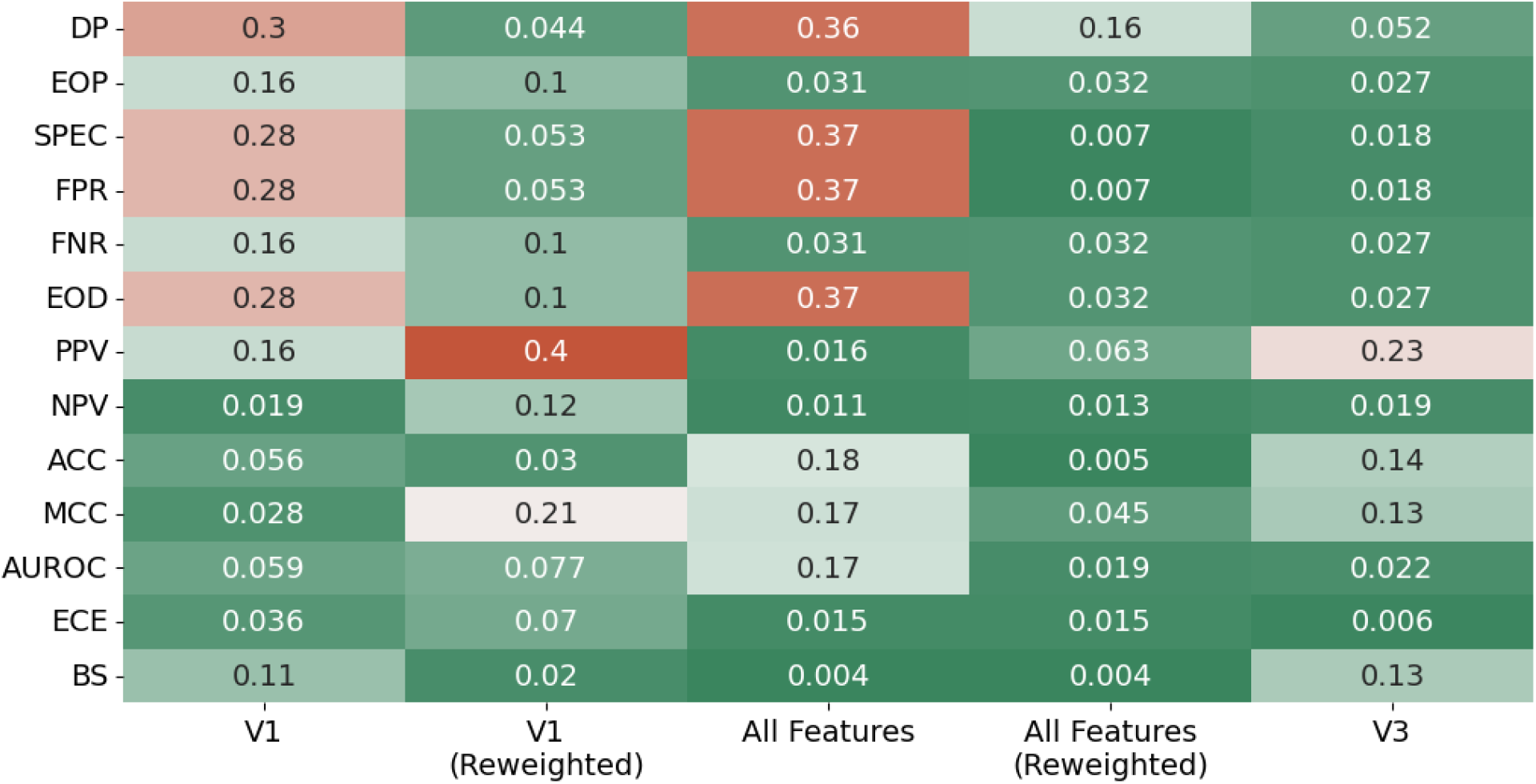
Fairness metric differences for the five models. Note that the best possible difference is zero (deep shade of green). V1 refers to UTICalc v1, V1 (Reweighted) refers to UTICalc v1 Reweighted, All Features refers to UTICalc v1 All Features, All Features (Reweighted) refers to UTICalc v1 All Features Reweighted, and V3 refers to UTICalc v3.

The disparity in sensitivity observed in UTICalc v1 is reflected in the EOP as a difference of 0.16, indicating that sensitivity differs between racial groups by approximately 16 percentage points (see Figure 1). Each of the four models improved the EOP (sensitivity parity) of UTICalc v1 in varying degrees, with the improvement ranging from 0.16 to 0.03. UTICalc v1 Reweighted showed improvement on all fairness metrics except for PPV, NPV, accuracy, and MCC.

UTICalc All Features showed improvement on half the fairness metrics, with no improvement on DP, Spec, FPR, EOD, Acc, MCC, and AUROC. While it was successful in reducing the EOP difference to 0.032, UTICalc v3 reduced the difference by a greater amount. Furthermore, the degree to which biases deteriorated was greater in UTICalc All Features compared to UTICalc v3.

UTICalc All Features Reweighted showed the greatest improvement across all fairness metrics compared to UTICalc v1. Even though it does not have the absolute smallest differences in the majority of metrics, it has small differences, with almost every difference below 0.07. Only two (DP and MCC) did not see improvement over UTICalc v1. Notably, it achieved this while having the highest AUROC, accuracy, and specificity of any model. The sensitivity decreased but remained above 90% in both racial groups (92.29% overall).

UTICalc v3 saw the best improvement in terms of EOP with a difference of just 0.027, but did not see improvement for PPV, NPV, accuracy, and MCC. Compared to UTICalc v1, all UTICalc v3 metrics demonstrating an improvement were statistically significant, with p-values less than 0.01. Surprisingly, UTICalc v3 decreased the Spec difference from 0.28 to 0.018. We attempted to improve UTICalc v3 by applying the exponentiated gradient reduction method. However, the reweighted model performed worse on all measures of fairness; hence, the results are not reported.

## Discussion

The reweighting of the features using the exponentiated gradient method showed promise by improving on UTICalc v1 and UTICalc All Features on the majority of the fairness metrics, although to a lesser extent than UTICalc v3. The exponentiated gradient method significantly reduced the difference in sensitivity between the two racial groups, but decreased the overall sensitivity of both reweighted models well below the target of 95%. This shows that when optimizing fairness of a model, it is essential to consider not only the magnitude of difference in the fairness metric of interest, but also monitor the overall performance. The model that included all features improved EOP, but at the cost of increasing disparities in several other fairness metrics. This suggests that the predictors not included in UTICalc v1 or UTICalc v3 result in different disparities that are not necessarily contained in the race predictor itself. Intriguingly, UTICalc v3 reveals some increased disparities in the discrimination metrics.

The observation that improvement in one fairness parity metric is associated with deterioration in other metrics, as seen in our results, is likely a consequence of the impossibility results described in Section xx which imply that only one of the three fairness metrics of DP, predictive parity (PPV parity and NPV parity) and EOD can hold simultaneously for a well-calibrated predictive model and a sensitive predictor like race that is susceptible to bias. This suggests that it may be impossible to reduce disparities in some discrimination metrics without increasing them in the others.

When compared to other models, the removal of race and its replacement with other predictors, as done in UTICalc v3, provided the greatest reduction in racial bias. While every model improved on EOP compared to UTICalc v1, they struggled to reduce disparities in other metrics. UTICalc v3 showed the greatest improvement in EOP, and most of that improvement can be attributed to a significant increase in sensitivity of the black group, without a corresponding decrease in the sensitivity of the nonblack group.

### Future work

We focused on optimizing fairness for better EOP using a single bias mitigation method, namely, the exponentiated gradient method. In the future, we plan to investigate additional mitigation methods for model optimization, as well as mitigation methods for other steps in the modeling process including preprocessing and postprocessing steps. We plan to investigate preprocessing optimization methods such as those described in [25] to increase fairness prior to model training. Due to UTICalc’s training data imbalance (roughly 25% black representation), preprocessing methods maybe particularly useful. Furthermore, we intend to investigate postprocessing methods like Threshold Optimizer [9], which can improve fairness by choosing a better probability threshold for predictions, or by selecting different probability thresholds for the different racial groups.

### Limitations

Our study has several limitations. As previously noted, we investigated a single bias mitigation method. The UTICalc dataset is imbalanced, containing only 25.2% black children compared to nonblack children, and we did not investigate modeling methods that are tailored to handle imbalanced datasets. Data imbalance across race can be a contributing factor to the remaining bias in model performance [26]. The dataset also has varying levels of missingness for each of the features, and since different models included different features and patients with missing data were excluded from model training, models were trained on varying sample sizes. The variance of the exponentiated gradient method may be affected by training the models on different sample sizes [23]. However, the percentage of black children in each of the training datasets was very similar, averaging around 25%. Our experiments were limited to the features that were originally collected for the development of UTICalc v1 and UTICalc v3. Additional features based on clinical data that are typically collected in the Emergency Department for febrile children suspected of having a UTI could potentially improve the models.

## Conclusions

Model respecifications that replaces a sensitive feature like race with new features or reweight features to optimize a fairness metrics, have the potential to reduce racial bias of models. Including all available features and reweighting those features as in UTICalc All Features Reweighted improved model fairness across many fairness metrics we considered. UTICalc v3 in which race was replaced by two new features but no reweighting was done performed the best on EOP. Further, our results show that, as predicted by previously described impossibility results, fairness cannot be simultaneously improved on all fairness metrics, and model respecification may improve racial fairness but decrease overall predictive performance. In future work, we will examine additional respecificaion methods that may further improve fairness of UTICalc All Features Reweighted.

## Data Availability

All data produced in the present study are available upon reasonable request to the authors.

## Acknowledgements

Research reported in this publication was supported by the National Institutes of Health under award number T15 LM007059 from the National Library of Medicine and under award number UL1 TR001857 from the National Center for Advancing Translational Sciences. It was also supported by a School of Computing and Information Predoctoral Fellowship to JWA.

